# Mpox vaccination coverage in the Democratic Republic of Congo: A systematic review and meta-analysis of uptake and acceptance (1970–2024)

**DOI:** 10.1101/2025.05.23.25327865

**Authors:** Fabrice Zobel Lekeumo Cheuyem, Adidja Amani, Adamu Abubakar Umar, Chabeja Achangwa, Brian Ngongheh Ajong, Solange Dabou, Jessy Goupeyou-Youmsi, Guy Roger Pilo Ndibo, Rick Tchamani, Ethel Ambo Eno, Jonathan Hangi Ndungo, Saralees Nadarajah

## Abstract

**Background:** The Democratic Republic of Congo (DRC) is recognized as the global epicenter of human Mpox. While vaccination is crucial for outbreak prevention, especially as the disease transitions from zoonotic spillover to sustained human-to-human transmission, comprehensive assessments of vaccination coverage trends across the country are notably absent from the literature. This systematic review and meta-analysis address this gap by providing the first pooled estimate of Mpox vaccine uptake and acceptance in the DRC over a 54-year period (1970-2024). Our study captures critical transitions, including the post-smallpox eradication era and recent global outbreaks, to identify temporal trends, geographic disparities in this high-risk setting.

**Methods:** We conducted this review following PRISMA guidelines, systematically searching PubMed, Scopus, ScienceDirect, Web of Sciences, CINAHL, and Embase. Grey literature was also searched to ensure comprehensiveness. Using random-effects models, we calculated pooled estimates for vaccine uptake and acceptance rates, with prespecified subgroup analyses examining variations by: (1) period, (2) geographic region, and (3) type of participants. We quantified heterogeneity using *I*^2^ statistics and conducted meta-regression to identify predictors of vaccination coverage heterogeneity. A p-value ⍰ 0.05 was considered statistically significant.

**Results:** Our analysis revealed a pooled Mpox vaccine uptake of 20.01% (95% CI: 7.45–43.75) with high heterogeneity (*I*^2^ = 99.4%, *p* < 0.001), indicating substantial variability across studies. Vaccine acceptance was higher at 54.17% (95% CI: 20.82–84.16) with high heterogeneity (*I*^2^ = 97.6%, *p* < 0.001). Temporal analysis showed a significant decline from 32.30% (95% CI: 14.62–57.75) coverage during 1970–2000 to 1.36% (95% CI: 0.29–6.11) in 2020–2024. Geographic disparities existed, with the Northwest regions achieving 47.11% (95% CI: 13.46–83.61) coverage compared to 5.47% (95% CI: 0.56–37.32) in Eastern conflict-affected zones. Meta-regression identified no significant predictors of coverage heterogeneity.

**Conclusion:** Despite moderate acceptance rates, actual Mpox vaccination uptake in the DRC remains critically low, with worsening coverage in recent years and substantial regional inequities. These findings underscore the urgent need for context-specific interventions to bridge the intention-action gap in this high-risk setting.

## Background

Mpox, previously termed monkeypox, is a zoonotic disease stemming from the monkeypox virus [1]. Before 2022, its presence was confined mainly to secluded rainforest communities in the western and central parts of Africa [2]. The virus spreads through direct contact with infected animals or humans, including sexual or skin-to-skin contact, respiratory droplets, and contaminated objects like clothing, towels, and bedding [3,4]. The most recent Mpox outbreak was initially declared in the UK on May 7, 2022, and the contagion has disseminated to other continents beyond Africa [5]. Subsequently, the disease outbreak was declared as a Public Health Emergency of International Concern on July 23, 2022 [6]. There are two types of Mpox, clade I and clade II. The ongoing global outbreak that started in 2022 is caused by clade II. The current epidemic in Eastern and Central Africa caused by clade I Mpox [7].

The Democratic Republic of Congo (DRC) has been the epicenter of human Mpox since the virus’s first identification in 1970 [2]. The current resurgence in the Democratic Republic of Congo (DRC) is linked to clade Ib, which appears to be spreading more rapidly through sexual contact and is associated with more virulent manifestations of the disease, especially in pediatric populations and immunodeficient patients [8,9]. Mpox demonstrates considerable disease severity and has caused significant mortality in the Democratic Republic of Congo [10]. To address this disease severity and save lives, the global health and scientific community has developed and deployed several vaccines for Mpox in the DRC, the disease’s most endemic focus worldwide [1,11].

Developing new vaccines for widespread use against Mpox and its emerging strains is a critical preventive strategy in the ongoing battle against this dynamic infection [12]. The smallpox vaccine has undergone three generations of technological advancements, with only the second and third versions now licensed. Three orthopoxviral vaccines are available: ACAM2000, JYNNEOS, and LC16m8 [12]. A substantial genetic homology exists between smallpox and Mpox, evidenced by shared immunological epitopes and markers. Owing to this close relationship, various studies indicate that smallpox vaccines confer robust cross-immunity against Mpox [13]. Furthermore, hypotheses have been advanced that the observed escalation in Mpox prevalence is attributable to a progressively immunologically naive global population, stemming directly from the cessation of smallpox immunization campaigns [14].

Despite four decades of Mpox surveillance, vaccination coverage remains poorly characterized across this vast area. Multiple studies reveal variable temporal trends, geographic disparities, and disparities in healthcare access, especially in conflict-affected regions [1,2,11,15–21]. The DRC’s health system - fragmented by decades of instability [5] - faces unique challenges in delivering vaccines to remote endemic zones like Equateur and Kivu [22]. Therefore, public health systems must take decisive action to prevent the global spread of Mpox, particularly among vulnerable groups [23]. This systematic review and meta-analysis on Mpox vaccination uptake and acceptance in Africa addresses critical evidence gaps in understanding immunization patterns across the continent’s highest-burden regions. As the Democratic Republic of Congo (DRC) reports over 80% of global Mpox cases, characterizing vaccine coverage dynamics becomes essential for outbreak response. There is a need for temporal trends following smallpox vaccination cessation, (2) geographic disparities exacerbated by conflict and healthcare access inequalities, and (3) the intention-action gap between vaccine acceptance and actual uptake. Our study addresses these knowledge gaps through the first comprehensive, half-century analysis of Mpox vaccination trends in the DRC. By synthesizing data from 1970 to 2024, we quantified temporal changes in coverage post-smallpox eradication, mapped regional inequities and assessed acceptance-uptake disparity. These insights will inform targeted vaccination strategies in this complex, high-priority setting.

## Methods

### Study design

This systematic review and meta-analysis examine the vaccination uptake and acceptance in the DRC between 1970 and 2024. The study was reported following the Preferred Reporting Items for Systematic Review and Meta-analysis (PRISMA) guidelines [24].

### Study setting

The DRC, the largest country in Sub-Saharan Africa, spans approximately 2,345,409 km^²^ [25]. It shares borders with nine countries (Angola, Burundi, the Central African Republic, the Republic of Congo, Rwanda, South Sudan, Tanzania, Uganda, and Zambia) 26]. The DRC comprises 26 provinces, including Kinshasa (the capital), Nord-Kivu, Sud-Kivu, Equateur, Tshuapa, and Bas-Huélé among others [10]. The health system is structured into three tiers: central, intermediate, and operational levels, with 519 health zones, 393 general referral hospitals, and 8,504 health areas delivering primary care [27]. With a population exceeding 105 million, the DRC has a median age of 16.7 years, a growth rate of 3.1%, and a life expectancy of 61.6 years [28]. The country faces recurrent infectious disease outbreaks, including Mpox, Ebola, Cholera, Measles and, Yellow fever, exacerbated by humanitarian crises linked to protracted civil unrest and armed conflict [29]. Socio-culturally, over 93% of the population identifies as Christian, with Catholics representing 42% [30]. These demographic, health, and geopolitical factors collectively shape the DRC’s unique challenges in disease surveillance and outbreak response [31].

### Eligibility criteria

#### Inclusion criteria

All observational studies addressing Mpox vaccine uptake and intention to vaccinate among general population, healthcare workers or other at-risk populations were included. No limitations were applied regarding language or time period. However, only those studies that were fully available, included sample size details, and presented relevant data on any aspect related to Mpox vaccine acceptance and uptake were included.

#### Exclusion criteria

Studies were deemed ineligible for inclusion if their research focus diverged from our investigative objectives or if their methodology differed from an observational design. Furthermore, incomplete articles were not considered, especially those characterized by insufficient data or an absence of the requisite results.

### Article searching strategy

Published research was identified through a systematic search on online databases, including PubMed, CINAHL, Embase, Web of Science, Scopus, ScienceDirect, and AJOL. The search process involved examining titles and abstracts using a keywords and Medical Subject Headings (MeSH). Boolean operators (“AND” and “OR”) were employed to refine the search as follow: “Mpox” OR “mpox” OR “Monkeypox” AND “vaccine” OR “vaccination” OR “acceptance” AND “DRC” OR “Democratic Republic of Congo” OR “Zaire” (Supplementary Table 1). A manual search for grey literature and additional publications not indexed in these databases was conducted in and Google Scholar (first 1000 entries assessed and no filter applied) to ensure comprehensive coverage. Furthermore, the reference lists of included studies were screened for relevant articles. The final search was completed on February 27, 2025.

### Data extraction

Data was extracted from all eligible articles using a pre-designed Microsoft Excel 2016 form to collect study characteristics. This form captured the first author’s name, study year, region, study design, study type, type of participant, setting, number of individuals willing to receive the vaccine, number of individuals who received the vaccine, and the sample size. Two authors independently assessed the relevance and quality of each article. Any disagreements between the reviewers were resolved through discussion with a third author to reach a consensus.

### Data quality assessment

The Joanna Briggs Institute (JBI) quality assessment tool was used to evaluate the quality of the studies included 32]. For cross-sectional studies, assessment criteria encompassed studies clear definition of inclusion criteria, comprehensive descriptions of study subjects and settings, validity and reliability of exposure measurements, use of objective, standardized criteria for outcome assessment, identification potential confounding factors, implementation of appropriate strategies to address them, and the appropriateness of statistical methods. For cohort study, this evaluation examined group comparability between exposed and unexposed populations, consistency and accuracy of exposure measurement, identification and adjustment for confounding factors, and confirmation of outcome-free status at baseline. The appraisal also considered validity of outcome measurement, adequacy and completeness of follow-up duration, strategies to address incomplete follow-up, and appropriateness of statistical analysis. Each criterion was scored as 1 (yes) or 0 (no or unclear). The overall risk of bias was categorized based on the proportion of criteria met as follow: low (>50%), moderate (>25-50%), or high (≤25%).

### Outcome measurement

The primary outcomes of this systematic review and meta-analysis were the Mpox vaccination uptake and the secondary outcome included the Mpox vaccine acceptance. The Mpox vaccine uptake and acceptance rate were calculated by dividing the number of people who received the vaccine and those who intended to receive it by the total number of participants.

### Operational definition

The vaccination uptake were measured in included studies base on the declaration of having received an Mpox vaccine or presented a Mpox vaccination card or a scar characteristic of the smallpox vaccine injection 15]. Meanwhile, the Mpox acceptance referred to the intention, defined as the unconditional willingness to receive the free Mpox vaccine if offered [11,33].

### Statistical analysis and synthesis

The Higgin’s *I*^2^ statistic was used to assess the heterogeneity between studies, categorizing heterogeneity as none (0%), low (<25%), moderate (25-75%), or high (>75%). Subgroup analyses were then conducted based on study period, region, setting, type of participants. Given the substantial heterogeneity observed across studies, reflecting divergent regional, temporal, and methodological contexts in Mpox vaccination data from the DRC, a random-effects model was selected to account for true variation in effect sizes beyond sampling error alone. This approach ensures that pooled estimates generalize to the broader population while conservatively weighting studies and acknowledging uncertainty introduced by underlying differences in study designs, populations, and implementation settings. Furthermore, meta-regression explored whether study characteristics including study year, location (region) and setting explained the variability in results. For subgroup analysis, the timeframe was divided into specific periods, including the smallpox vaccine era, 2010–2019: limited Mpox vaccine interest; 2020–2024: increased focus due to global outbreaks. The regions were grouped into geographical specific group including the Northwest endemic zone (historically high Mpox transmission zone encompassed Equateur, Tshuapa and Bas-Uélé); the Central/South endemic zone (moderate Mpox activity comprised Sankuru and Kasai Oriental); and the Easthern conflict affected zone (lower Mpox activity but high population movement included South Kivu, North Kivu and Tshopo). Generalized Linear Mixed Models (GLMM), coupled with the Probit-Logit Transformation (PLOGIT) were used as they are ideal for meta-analyses of binary or proportion data as they directly model binomial outcomes, effectively account for between-study heterogeneity using random effects, and inherently handle studies with 0% or 100% events without requiring continuity corrections 34]. Statistical significance was set at a p-value of <0.05. All analyses were performed using the ‘meta’ package in R Statistics version [4.4.2 35].

### Publication bias and sensitivity test

Publication bias was assessed visually using a funnel plot. The symmetry of the inverted funnel shape suggested the absence of publication bias. The Egger’s and Begg’s tests were also performed to test the significance any visually assessed publication bias. Sensitivity analysis was conducted by iteratively excluding one study at a time to assess the robustness of the findings. This tested the robustness of our primary meta-analysis results to potential outliers, particularly given the high heterogeneity potential. By iteratively excluding each study, we aimed to quantify how removal of specific studies (e.g., those with extreme proportions or large sample sizes) altered the pooled estimate and determine if conclusions remained consistent across methodological variations.

## Results

A total of 1,845 entries were obtained through an online database search (n = 1,796) and a manual search (n = 49). After removing 167 redundant entries, 1,678 distinct entries underwent title and abstract review, followed 8 by a complete text evaluation for suitability. Ultimately, 18 reports satisfied the inclusion criteria and were incorporated into the meta-analysis (Fig. 1).

**Fig. 1.**
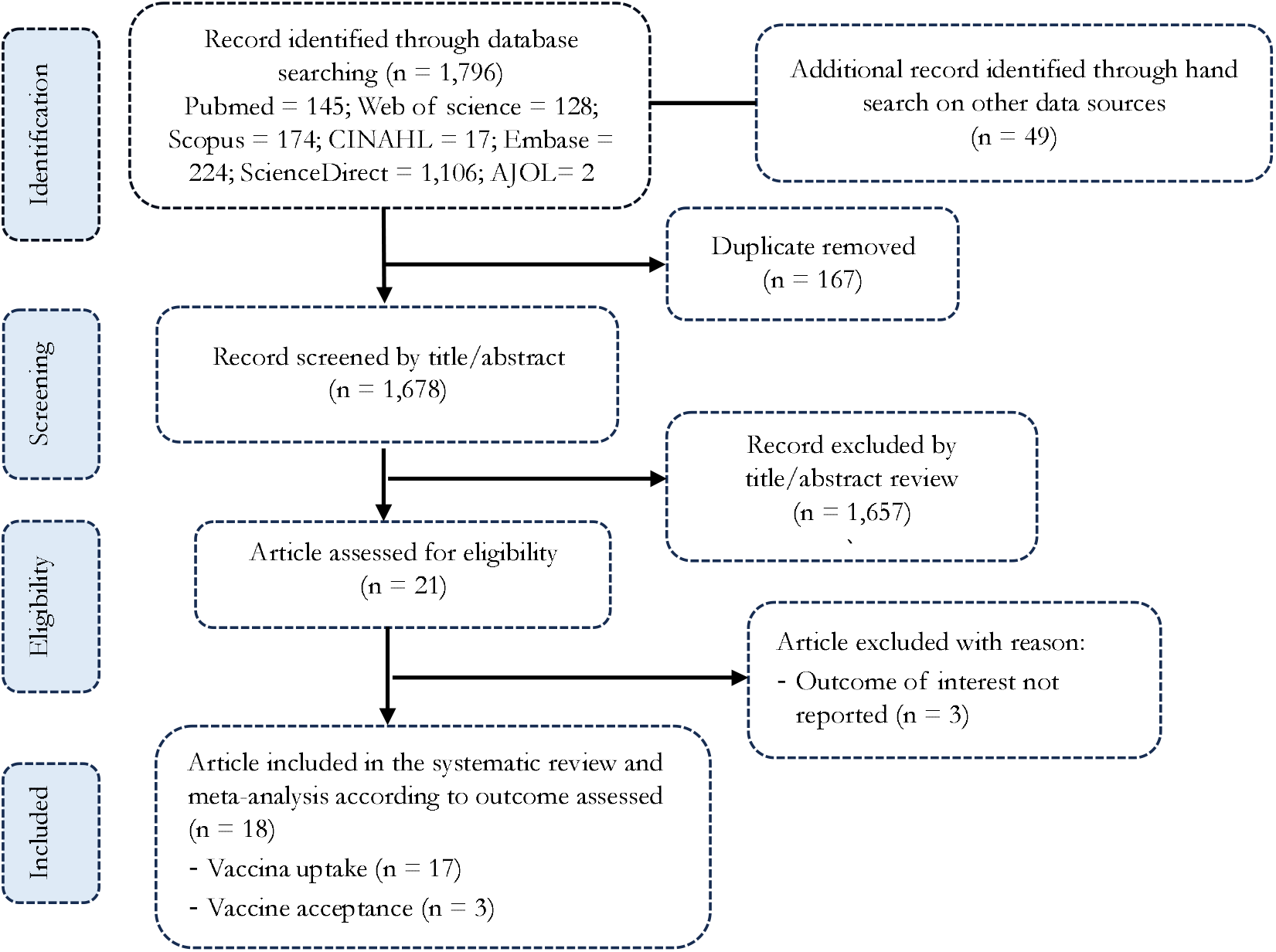
PRISMA diagram flow of studies included in the meta-analysis.

### Studies selection

### Characteristics of studies included

Eighteen studies carried out in community and hospital settings across the DRC between 1970 and 2024, were included in this comprehensive analysis 1,2,11,15–21,36–43]. Most of these studies (n = 15) employed surveillance and investigation for data collection, focusing on the general population and healthcare workers to describe Mpox vaccination acceptance and uptake. Most of the studies (n = 16) presented a low risk of bias (Table 1).

**Table 1.** Characteristic of studies assessing Mpox vaccination in DRC, 1970-2024.

| Author | Study Year<sup>1</sup> | Region | Setting | Study population | Study type | Sampling | Risk of bias | Outcome of interest | Sample size vaccination survey | Summary of findings |
| --- | --- | --- | --- | --- | --- | --- | --- | --- | --- | --- |
| Ladnyj *et al.* [15] | 1970 | Equateur | Community | General population | Cross-sectional | Non-probabilistic | Low | Vaccine uptake | 1132 | The unvaccinated young patient's relatives and most villagers showed signs of smallpox vaccination or previous infection. |
| Breman *et al.* [2] | 1979 | Equateur | Community | General population | Cross-sectional | Non-probabilistic | Low | Vaccine uptake | 38 | Of the 47 reported cases in Central and West Africa, 38, representing the vast majority, occurred in Zaire. These cases, largely in children under 10, presented with a 17% case-fatality rate. |
| Mutombo *et al.* [36] | 1982 | South Kivu | Community | General population | Cross-sectional | Non-probabilistic | Low | Vaccine uptake | 1367 | The rash's onset was consistent with chimpanzee transmission, but the primary monkeypox reservoir remained unknown. |
| Jezek *et al.* [18] | 1984 | Nationwide | Community | General population | Cross-sectional | Non-probabilistic | Low | Vaccine uptake | 2083 | A study of 2,510 contacts of monkeypox patients revealed a significant number of secondary cases, primarily in children, with waning immunity observed in vaccinated individuals, though vaccination still offered considerable protection. |
| Jezek *et al.* [37] | 1985 | Nationwide | Community | General population | Cross-sectional | Non-probabilistic | Low | Vaccine uptake | 282 | unvaccinated individuals, particularly young children, experienced an 11-15% case-fatality rate, while vaccinated individuals showed no deaths and altered disease presentation. |
| Mwamba *et al.* [38] | 1997 | Kasai Oriental | Community | General population | Cross-sectional | Non-probabilistic | Low | Vaccine uptake | 84 | The 1996-97 investigation suggested most monkeypox cases resulted from person-to-person transmission, a shift from earlier reports of primarily animal-to-human spread. |
| Hutin *et al.* [39] | 1997 | Sankuru | Community | General population | Cross-sectional | Non-probabilistic | Low | Vaccine uptake | 84 | This outbreak presented with notable attack and case-fatality rates. The cessation of smallpox vaccination in 1983, following global eradication, contributed to an increased population susceptibility to monkeypox. |
| Aplogan *et al.* [40] | 1997 | Kasai Oriental | Community | General population | Cross-sectional | Non-probabilistic | Low | Vaccine uptake | 339 | There was a high prevalence of Mpox among children and a mix of primary and secondary transmission patterns across numerous villages. The outbreak was localized outbreaks and travel, close contact within households and neighborhoods facilitated the secondary case transmission. |
| Rimoin *et al.* [41] | 2007 | Sankuru | Community | General population | Cross-sectional | Non-probabilistic | Low | Vaccine uptake | 760 | This study revealed a dramatic increase in cases, likely due to the cessation of smallpox vaccination and the subsequent rise in a immunologically naive population. |
| Nolen *et al.* [42] | 2012 | Tshuapa | Hospital | General population and healthcare worker | Cross-sectional | Non-probabilistic | Low | Vaccine acceptance and uptake | 148 | Half of possible cases were lab-confirmed monkeypox. 50% household attack rate, multiple family transmissions, and a mean 8-day incubation period (4-14 days) were observed. |
| Petersen *et al.* [1] | 2014 | Tshuapa | Community | Healthcare worker | Cross-sectional | Non-probabilistic | Low | Vaccine uptake and acceptance | 14 | These findings suggest that while a smallpox vaccination scar may offer some protection against monkeypox, healthcare workers remain at risk of infection. The presence of co-infections further complicates the picture. |
| McCollum *et al.* [19] | 2014 | Kivu (North and South) | Community | General population | Cross-sectional | Non-probabilistic | Low | Vaccine uptake | 30 | Report of the first confirmed cases of monkeypox (Mpox) in forested areas of North and South Kivu Provinces, which aligns with ecological predictions for suitable Mpox transmission zones. |
| Whitehouse *et al.* [20] | 2014 | Tshuapa | Community | General population | Cross-sectional | Non-probabilistic | Low | Vaccine uptake | 1057 | Increased incidence compared to previous decades, likely due to waning smallpox immunity. While males generally had higher infection rates, females reported frequent contact with symptomatic individuals. Animal exposures were most common in males. |
| Mande *et al.* [43] | 2019 | Bas-Uélé | Community | General population | Cross-sectional | Non-probabilistic | Low | Vaccine uptake | 71 | Among 77 suspected cases, PCR revealed 27.3% monkeypox, 58.4% chickenpox, and 14.3% negative. Monkeypox cases showed distinct skin lesions. |
| Brosius *et al.*<br>[16] | 2024 | South Kivu | Hospital | General<br>population | Cohort | Non-probabilistic | Moderate | Vaccine<br>uptake | 308 | Most suspected cases were PCR-positive for Mpox. Most cases reported contact with known Mpox, primarily spouses/partners in adults and family in children. Genital lesions were common in adults. Hospitalized mortality was low. |
| Petrichko *et al.*<br>[11] | 2024 | Nationwide | Community | General<br>population | Cross-sectional | Non-probabilistic | Moderate | Vaccine<br>acceptance | 5226 | In the DRC, 61% accepted a Mpox vaccine, with higher rates among healthcare workers and in endemic regions. |
| Kombozi *et al.*<br>[21] | 2024 | Tshopo | Community | General<br>population | Cross-sectional | Probabilistic | Low | Vaccine<br>uptake | 460 | Occupational activities, such as hunting and agriculture, alongside economic vulnerability and direct exposure to infected animals or individuals, play a significant role in Mpox transmission. These results underscore the complex interplay between human behavior, environmental interaction, and disease spread. |
| Vakaniaki *et al.*<br>[17] | 2024 | South Kivu | Community | General<br>population | Cross-sectional | Non-probabilistic | Low | Vaccine<br>uptake | 241 | The Mpox outbreak in eastern DRC was caused by a distinct Clade I Mpox lineage, differing from historical zoonotic patterns. The outbreak, predominantly affected young adults including a significant proportion of female sex workers, suggests a shift towards human-to-human transmission, potentially involving sexual contact. |

<sup>1</sup> Date of study completion; HCW: Healthcare worker; DRC: Democratic Republic of Congo; PCR: Polymerase chain reaction

**Table 2.** Subgroup analysis of Mpox vaccination coverage in DRC, 1970-2024.

| Subgroup | Sample size | Coverage (%) | 95% CI limits |  | Number of studies | Heterogeneity statistic<sup>1</sup> |  |
| --- | --- | --- | --- | --- | --- | --- | --- |
|  |  |  | Lower | Upper |  | *I*<sup>2</sup>(%) | *p*-value |
| **Study period<sup>2</sup>** |  |  |  |  |  |  |  |
| 1970-2000 | 5409 | 32.30 | 10.41 | 66.20 | 8 | 99.6 | ≪0.001 |
| 2001-2020 | 2080 | 32.89 | 9.64 | 69.24 | 6 | 97.6 | ≪0.001 |
| 2021-2024 | 1009 | 1.36 | 0.29 | 6.11 | 3 | 0.0 | 0.635 |
| **Study period<sup>3</sup>** |  |  |  |  |  |  |  |
| ≪ 2022 | 7489 | 32.60 | 14.62 | 57.75 | 14 | 99.5 | ≪0.001 |
| 2022+ | 1009 | 1.36 | 0.29 | 6.11 | 3 | 0.0 | 0.635 |
| **Region** |  |  |  |  |  |  |  |
| Equateur | 1170 | 53.80 | 2.74 | 97.97 | 2 | 98.6 | ≪0.001 |
| Kasai Oriental | 423 | 10.01 | 4.48 | 20.87 | 2 | 91.3 | 0.007 |
| Sankuru | 844 | 7.42 | 2.71 | 18.70 | 2 | 94.6 | 0.004 |
| Tshuapa | 1219 | 20.87 | 8.75 | 42.04 | 3 | 94.2 | ≪0.001 |
| Kivu (North and South) | 1946 | 5.58 | 0.26 | 57.30 | 4 | 96.0 | ≪0.001 |
| Bas-Uélé | 71 | 92.96 | 84.33 | 97.67 | 1 | - | - |
| Tshopo | 460 | 3.48 | 2.00 | 5.59 | 1 | - | - |
| Nationwide | 2365 | 53.36 | 5.16 | 96.00 | 2 | 99.8 | ≪0.001 |
| **Zone<sup>4</sup>** |  |  |  |  |  |  |  |
| Northwest endemic zone | 2460 | 47.11 | 13.46 | 83.61 | 6 | 99.5 | ≪0.001 |
| Central/South endemic zone | 1267 | 8.63 | 4.46 | 16.04 | 4 | 91.3 | ≪0.001 |
| Eastern conflict-affected zone | 2406 | 5.47 | 0.56 | 37.32 | 5 | 97.9 | ≪0.001 |
| Nationwide | 2365 | 53.36 | 5.16 | 96.00 | 2 | 99.8 | ≪0.001 |
| **Setting** |  |  |  |  |  |  |  |
| Community | 8190 | 22.59 | 8.37 | 48.35 | 16 | 99.5 | ≪0.001 |
| Hospital | 308 | 2.27 | 0.92 | 4.63 | 1 | - | - |
| **Participant** |  |  |  |  |  |  |  |
| General population | 8336 | 18.37 | 5.86 | 44.82 | 15 | 99.5 | ≪0.001 |
| General population and Healthcare worker | 148 | 21.61 | 15.28 | 29.13 | 1 | - | - |
| Healthcare worker | 14 | 50.00 | 23.04 | 76.96 | 1 | - | - |

<sup>1</sup> Random effects model; CI: Confidence interval; <sup>2</sup> 1970–2000: smallpox vaccine era; 2001–2020: Limited Mpox vaccine interest; 2021–2024: Increased
focus due to global outbreaks;<sup>3</sup> Period before and after the global outbreak declaration;<sup>4</sup> Northwest endemic zone (historically high Mpox transmission zone) = Equateur, Tshuapa and Bas-Uélé; Central/South endemic zone (moderate Mpox activity) = Sankuru and Kasai Oriental; Eastern conflict affected zone (lower Mpox activity but high population movement) = South Kivu, North Kivu and Tshopo.

### Mpox vaccine uptake

A random effects meta-analysis of 17 studies (n = 8498 participants) estimated an overall Mpox vaccination uptake rate of 20.01% (95% CI: 7.45–43.75), with substantial heterogeneity (*I*^2^=99.4%, *p* < 0.001). Study-specific rates ranged from 0% to 94.08% (Fig. 2).

**Fig. 2.**
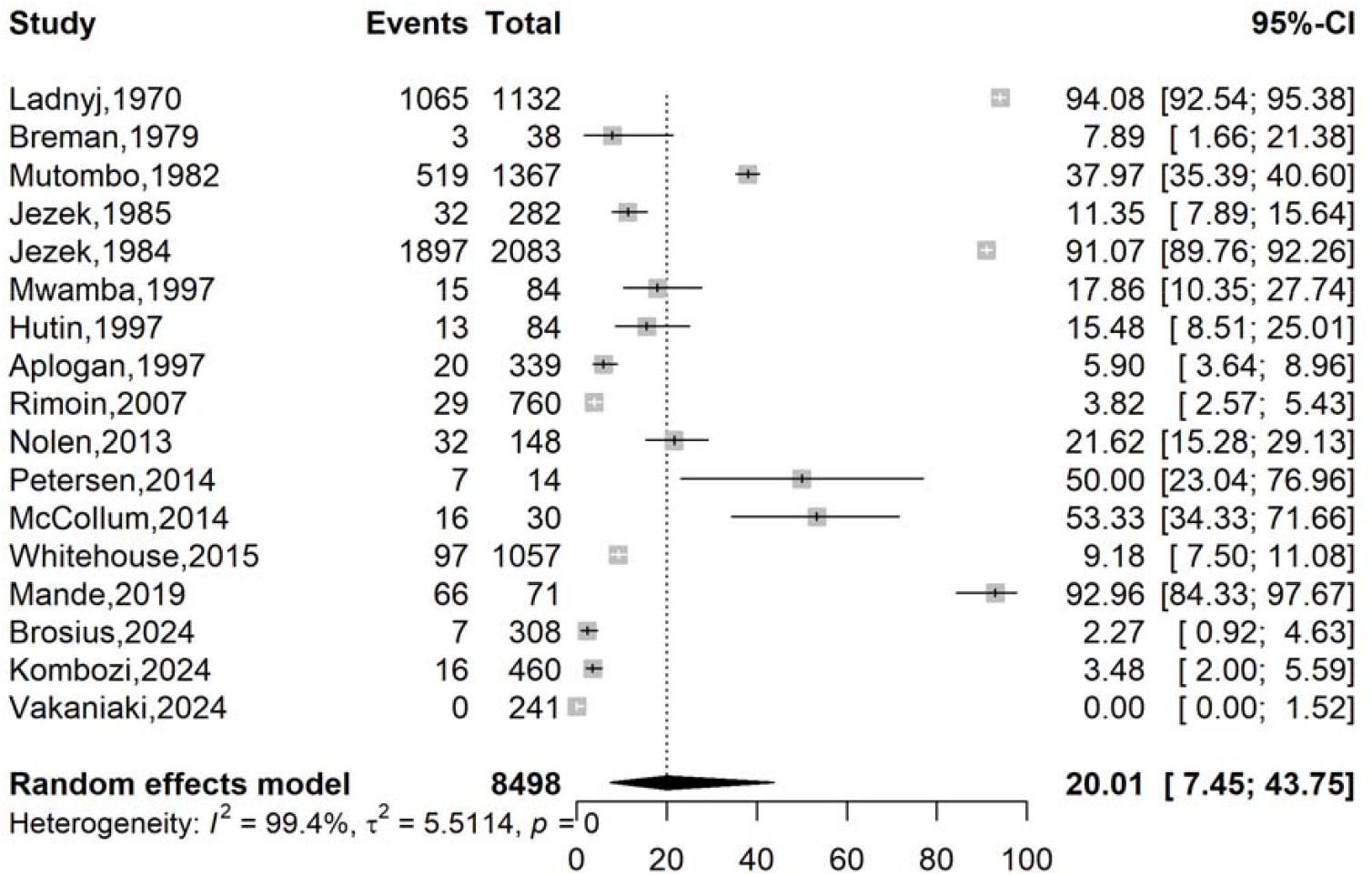
Pooled Mpox vaccination coverage estimate in DRC, 1970-2024.

### Mpox vaccine acceptance

A random effects meta-analysis of three studies (n = 5,388) yielded a pooled vaccine acceptance rate of 54.17% (95% CI: 20.82–84.16), with high heterogeneity (*I*^2^=97.6%, *p* < 0.001). Individual study rates varied widely, from 18.92% (Nolen *et al*., 2013) [42] to 85.71% (Petersen, et al., 2014) [1], with the largest study (Petrichko *et al*., 2024) [11] reporting 61.00% (95% CI: 59.66–62.33) (Fig. 3).

**Fig. 3.**
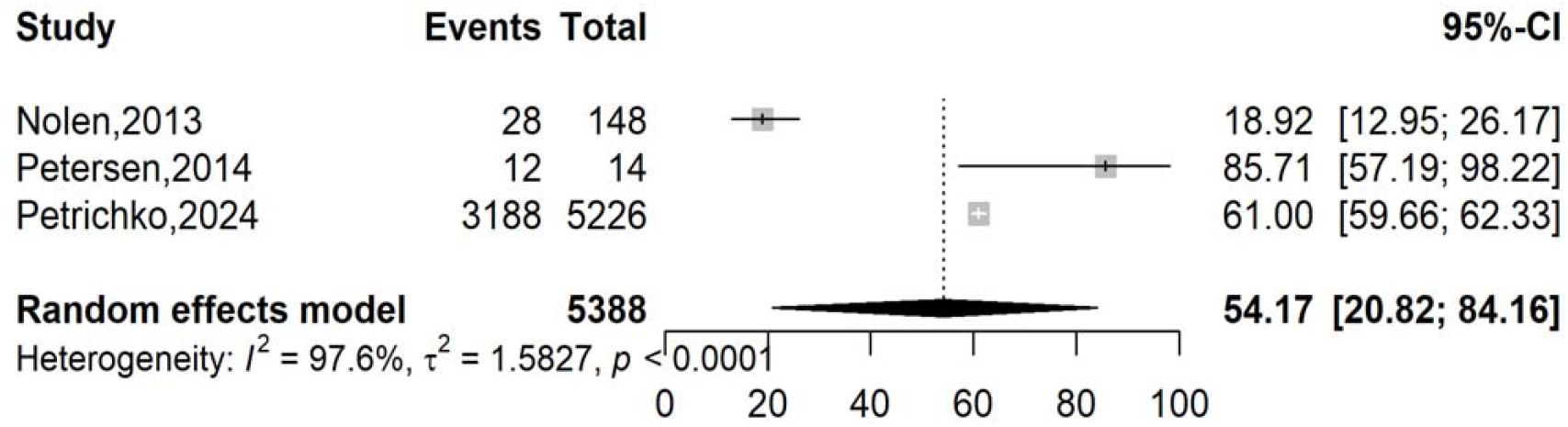
Pooled Mpox vaccination acceptance estimates in DRC, 1970-2024.

### Subgroup analysis

The subgroup analysis revealed significant disparities in Mpox vaccination coverage across different periods, regions, and study designs in the DRC (1970–2024). The vaccination coverage was highest in earlier decades (32.30% in 1970–2000; 32.89% in 2001–2020) but decreased to 1.36% during 2021–2024, suggesting recent declines significantly (*p* = 0.001) in vaccination efforts. Geographically, Equateur (53.60%, n = 2 studies) and Bas-Uélé (92.96%, n = 1 study) had notably high coverage, while Sankuru (7.42%) and Tshopo (3.48%) lagged. Nationwide studies reported highly variable estimates (51.23%, 95% CI: 0–100%, n = 2 studies).

### Meta-regression analysis

The meta-regression analysis found no statistically significant predictors of Mpox vaccination coverage heterogeneity in the DRC (1970–2024). While the study year showed a marginal negative trend in univariate analysis (β = −0.0545, *p* = 0.078), suggesting a possible decline in coverage over time, this association was not significant after adjustment (β = −0.0427, *p* = 0.248). Similarly, region and setting (hospital vs. community) demonstrated no significant effects in either univariate or multivariate models (p > 0.1 for all) (Table 3).

**Table 3.** Meta-regression analysis of Mpox vaccination coverage and acceptance in DRC, 1970-2024.

| Estimate | Moderator | Univariate |  | Multivariate |  |
| --- | --- | --- | --- | --- | --- |
| | | Unadjusted coefficient ($\beta$) | *p*-value | Adjusted coefficient ($\beta$) | *p*-value |
| Vaccination uptake |  |  |  |  |  |
| Study year | Year | -0.0545 | 0.078 | -0.0427 | 0.248 |
| Region | Region vs. Nationwide | -1.6282 | 0.341 | -0.7019 | 0.700 |
| Setting | Hospital vs. Community | -2.6276 | 0.258 | -1.5533 | 0.530 |

### Publication bias

While the left shift of the funnel plot suggested potential publication bias, Egger’s test (*p* = 0.203) and Begg’s test (*p* = 0.249) did not indicate any statistically significant bias (Fig. 4).

**Fig. 4.**
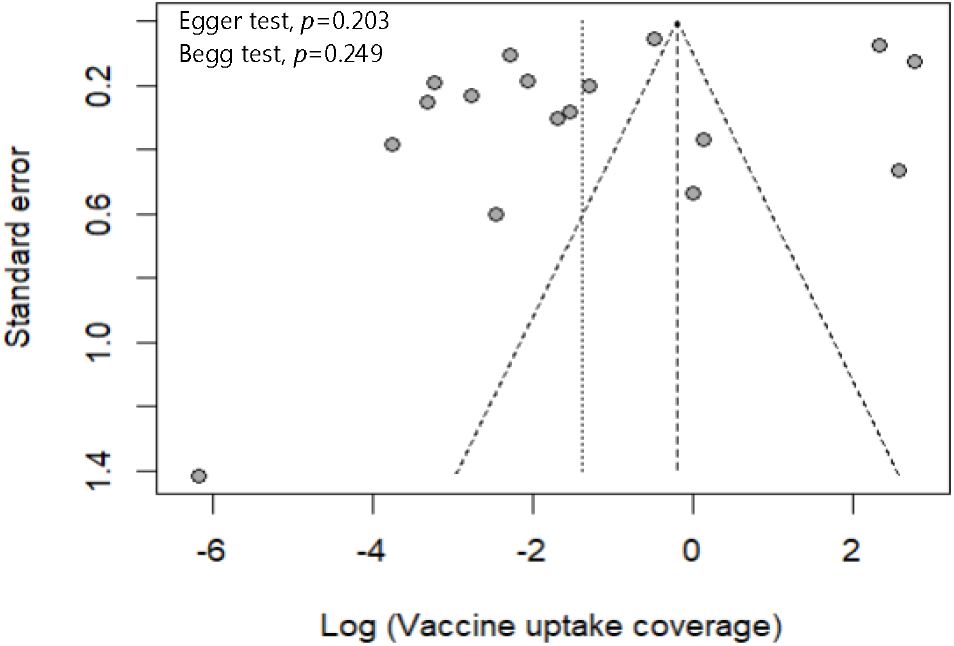
Funnel plot with pseudo 95% confidence limits and tests assessing the publication bias of studies included.

### Sensitivity test analysis

Sensitivity analysis indicated that no single study significantly influences the pooled Mpox vaccination coverage estimate reflecting the robustness of the findings (Fig. 5).

**Fig. 5.**
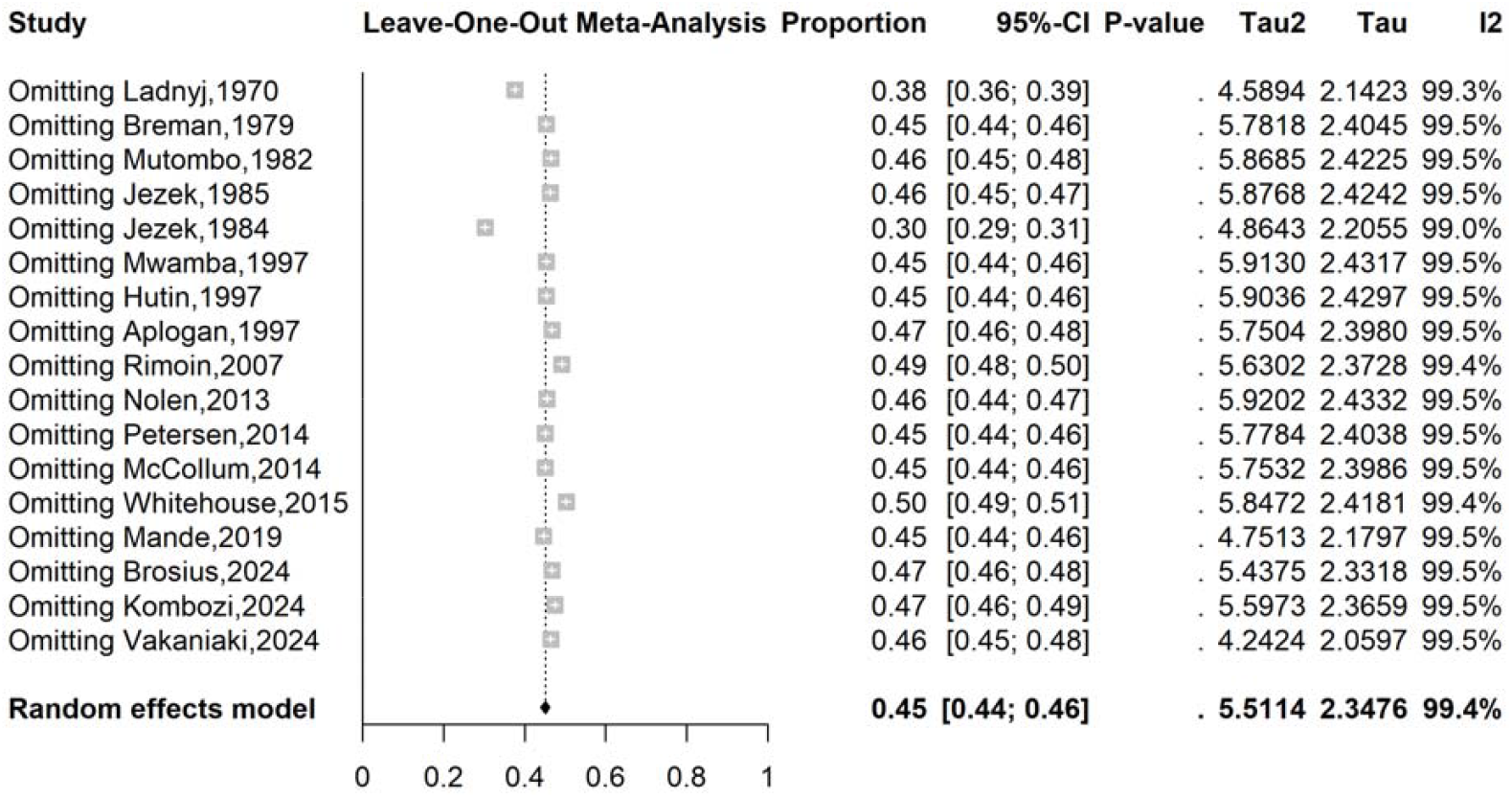
Sensitivity analysis of the pooled Mpox vaccination coverage in DRC, 1970-2024.

## Discussion

This systematic review and meta-analysis were primarily conducted to determine the Mpox vaccine uptake rate in the world’s most endemic focus. It synthesized data from 18 studies to provide a comprehensive overview of the DRC from 1970 to 2024.

### Vaccine uptake

This meta-analysis comprehensively assesses Mpox vaccination coverage in the DRC from 1970 to 2024, revealing critical insights into its temporal trends, regional disparities, and methodological challenges. The pooled estimate (20.01%, 95% CI: 7.45–43.75) and subgroup analyses demonstrated high heterogeneity (*I*^2^ > 99.4%), reflecting wide variability in coverage across regions, periods, and study designs. Our results corroborated findings from a global meta-analysis of Mpox vaccine uptake, which revealed a pooled rate of 30.9% (95% CI: 21.0–41.7), but this was higher than that of the WHO African region at 5.0% (95% CI: 3.7–6.7) 33]. This highlights a substantial disparity in vaccine coverage between the African region and the global average. This vaccination profile could be attributed to structural barriers, including limited healthcare infrastructure, cold-chain challenges, and inequitable vaccine distribution in Africa, all of which reduce access compared to high-income regions with robust systems. Additionally, socioeconomic factors such as lower health literacy, vaccine hesitancy driven by mistrust, and competing health priorities further widen the gap in coverage [44,45].

Our analysis revealed a notable temporal trend in Mpox vaccination coverage within the DRC. Coverage was substantially higher in studies conducted between 1970 and 2000, with a pooled estimate of 32.30% (95% CI: 10.41-66.20). However, this figure plummeted to a mere 1.36% in studies conducted post-2020. This represents a significant reduction of over 95% in estimated vaccination coverage, indicating a critical shift in the landscape of Mpox prevention efforts. This decline may be associated with challenges in vaccine supply chains, reduced access to healthcare facilities, and a diversion of resources to outbreak response [46,47]. This likely contributed to the substantial decrease in routine vaccination activities. The significant drop in Mpox vaccination coverage post-2020 seriously affects public health in the DRC. Reduced population immunity increases the risk of future outbreaks, potentially with greater severity and wider spread, particularly if healthcare systems remain strained [10,48].

To explore potential geographical variations in Mpox vaccination coverage, we conducted a stratified analysis across three distinct zones within the DRC: the Northwest endemic zone, the Central/South endemic zone, and the Eastern conflict-affected zone. Our study revealed significant disparities in pooled estimates across these regions. The Northwest endemic zone exhibited the highest estimated coverage/acceptance at 47.11% (95% CI: 13.46-83.61), while the Eastern conflict-affected zone showed a considerably lower estimate of 5.47 (95 CI: 0.56-37.32). The higher estimated coverage in the Northwest endemic zone could be attributed to the region’s more extended history with Mpox outbreaks, potentially leading to greater community awareness and established vaccination programs over time [2,15]. The lower coverage observed in the Central/South endemic zone, or the Eastern conflict-affected zone, likely reflects differing healthcare priorities or access challenges compared to the Northwest. The complex interplay of conflict-related disruptions to healthcare access alongside potential targeted interventions by humanitarian organizations may also explain these findings [16,19].

Future research should prioritize comprehensive investigations into the intricate interplay of healthcare infrastructure, community engagement strategies, and the pervasive impact of conflict on vaccination efforts across different regions. By meticulously dissecting these nuances, we can pave the way for developing and implementing targeted, equitable, and ultimately more effective strategies to strengthen immunization programs in DRC.

### Vaccine acceptance

Our study’s pooled Mpox vaccine acceptance rate was 54.17% (95% CI: 20.82-84.16), consistent with a global meta-analysis and an African multinational study reporting acceptance rates from 51.1% to 74.4% [5,33]. However, we identified a substantial discrepancy between vaccine acceptance (54.17%) and actual uptake (20.01%), revealing a critical implementation gap in Mpox vaccination programs. This disparity likely results from multiple factors, including temporal variations in study periods and uneven vaccine availability across different settings.

Comprehensive vaccination strategies are urgently needed to address this gap and convert vaccination intentions into actual coverage [11,49]. Such efforts are vital as individuals who initially intend to vaccinate but remain unvaccinated are at high risk of vaccine hesitancy or refusal over time [50,51]. Therefore, effective public health interventions must extend beyond promoting acceptance to include practical measures that improve vaccine accessibility, overcome logistical challenges, and strengthen community trust in vaccination programs.

Closing this implementation gap is especially crucial given Mpox’s severe health consequences in the DRC, where vulnerable populations - including children and immuno-compromised individuals - experience high rates of morbidity and mortality. The clinical management of Mpox is further complicated by frequent coinfections with varicella-zoster virus, underscoring the urgent need for effective vaccination programs [10,52].

## Strength and limitations

This systematic review and meta-analysis provide a comprehensive synthesis of Mpox vaccination trends in the DRC over five decades, offering valuable insights into temporal patterns, geographic disparities, and implementation gaps through robust analysis of 18 diverse studies. However, this review has several limitations. The small number of included studies for Mpox vaccine acceptance (□3). Additionally, the small sample sizes in some subgroup analyses resulted in wide confidence intervals, limiting the precision of the findings. The predominance of non-probabilistic surveillance data might limit the inference. In addition, the vaccination status relied primarily on individual reports or the presence of a scar of vaccination instead of the presentation of a vaccination card testifying to the reception of the vaccine. Due to small number of studies assessing Mpox vaccine acceptance in DRC, further statistical analyses were not performed. Despite these limitations,

## Conclusions

This systematic review revealed critical findings about Mpox vaccination in the DRC. A dramatic decline in coverage from 1970 to 2024 signals systemic failures in maintaining population immunity, particularly after the global strategy to eradicate smallpox. The stark disparity between vaccine acceptance and actual uptake highlights substantial implementation gaps beyond willingness. Geographic inequities, with Northwest zones achieving higher coverage than Central and armed conflict regions, underscore how conflict and healthcare access shape vaccination outcomes. Two main priorities identified include targeted campaigns for underserved areas like Sankuru and Tshopo provinces to convert acceptance into uptake through community-engaged delivery models. As Mpox continues to evolve and poses a public health threat in this endemic focus, these results provide both a warning and a roadmap to close the vaccination gaps before the outbreak escalates beyond control. Future success will require integrating these empirical findings with local knowledge to build equitable, sustainable protection for all communities.

## Data Availability

The sources of data supporting this systematic review are available in the reference. All data generated or analyzed during this study are included in this published article and supplemental material.

## Abbreviations

CI: Confidence interval
DRC: Democratic Republic of Congo
HCW: Healthcare worker
MeSH: Medical subject headings
Mpox: Monkeypox
PRISMA: Preferred reporting items for systematic reviews and meta-analysis

## Declarations

### Ethical approval and consent to participate

Not applicable.

### Consent for publication

Not applicable.

### Competing interests

All authors declare no conflicts of interest and have approved the final version of the article.

### Funding source

This research did not receive any specific grant from funding agencies in the public, commercial or not-for-profit sectors.

### Author contributions

F.Z.L.C. conceived the original idea of the study. F.Z.L.C and C.A. conducted the literature search. F.Z.L.C., C.A. and A.A. selected the studies, extracted the relevant information, and synthesized the data. F.Z.L.C., A.A.U. and S.N. performed the analyses. F.Z.L.C. wrote the first draft of the manuscript. All authors critically reviewed and revised successive drafts of the manuscript. All authors read and approved the final manuscript.

## Acknowledgements

None.

## Notes

### Competing Interest Statement

The authors have declared no competing interest.

### Author Declarations

All data sources are available in the reference list.

### Summary of Updates

Searching was conducted in additional databases. The statistics were changed to be more accurate, and the whole manuscript has been revised accordingly.

