## Additional File for "Mpox vaccination coverage in the Democratic Republic of Congo: A systematic review and meta-analysis of uptake and acceptance (1970–2024)"

Supplementary Table 1 Searching Strategies for online databases

| Database | Search Term | Results |
| --- | --- | --- |
| PubMed | ((Mpox) OR (mpox) OR (Monkeypox)) AND ((vaccine) OR (vaccination) OR (acceptance)) AND ((DRC) OR (Democratic Republic of Congo) OR (Zaire)) | 145 |
| Web of Science | ALL=((Mpox) OR (mpox) OR (Monkeypox)) AND ((vaccine) OR (vaccination) OR (acceptance)) AND ((DRC) OR (Democratic Republic of Congo) OR (Zaire)) | 128 |
| Scopus | TITLE-ABS-KEY((Mpox) OR (mpox) OR (Monkeypox)) AND ((vaccine) OR (vaccination) OR (acceptance)) AND ((DRC) OR (Democratic Republic of Congo) OR (Zaire)) | 174 |
| CINAHL | ((Mpox) OR (mpox) OR (Monkeypox)) AND ((vaccine) OR (vaccination) OR (acceptance)) AND ((DRC) OR (Democratic Republic of Congo) OR (Zaire)) | 17 |
| Embase | ((Mpox) OR (mpox) OR (Monkeypox)) AND ((vaccine) OR (vaccination) OR (acceptance)) AND ((DRC) OR (Democratic Republic of Congo) OR (Zaire)) | 224 |
| ScienceDirect | ((Mpox) OR (mpox) OR (Monkeypox)) AND ((vaccine) OR (vaccination) OR (acceptance)) AND ((DRC) OR (Democratic Republic of Congo) OR (Zaire)) | 1,106 |
| African Journals Online (AJOL) | Mpox OR mpox OR Monkeypox AND vaccine OR vaccination OR acceptance AND DRC OR Democratic Republic of Congo OR Zaire | 2 |

**Vaccination coverage**

Period

**Event rate (%)**

**Vaccine coverage (%)**

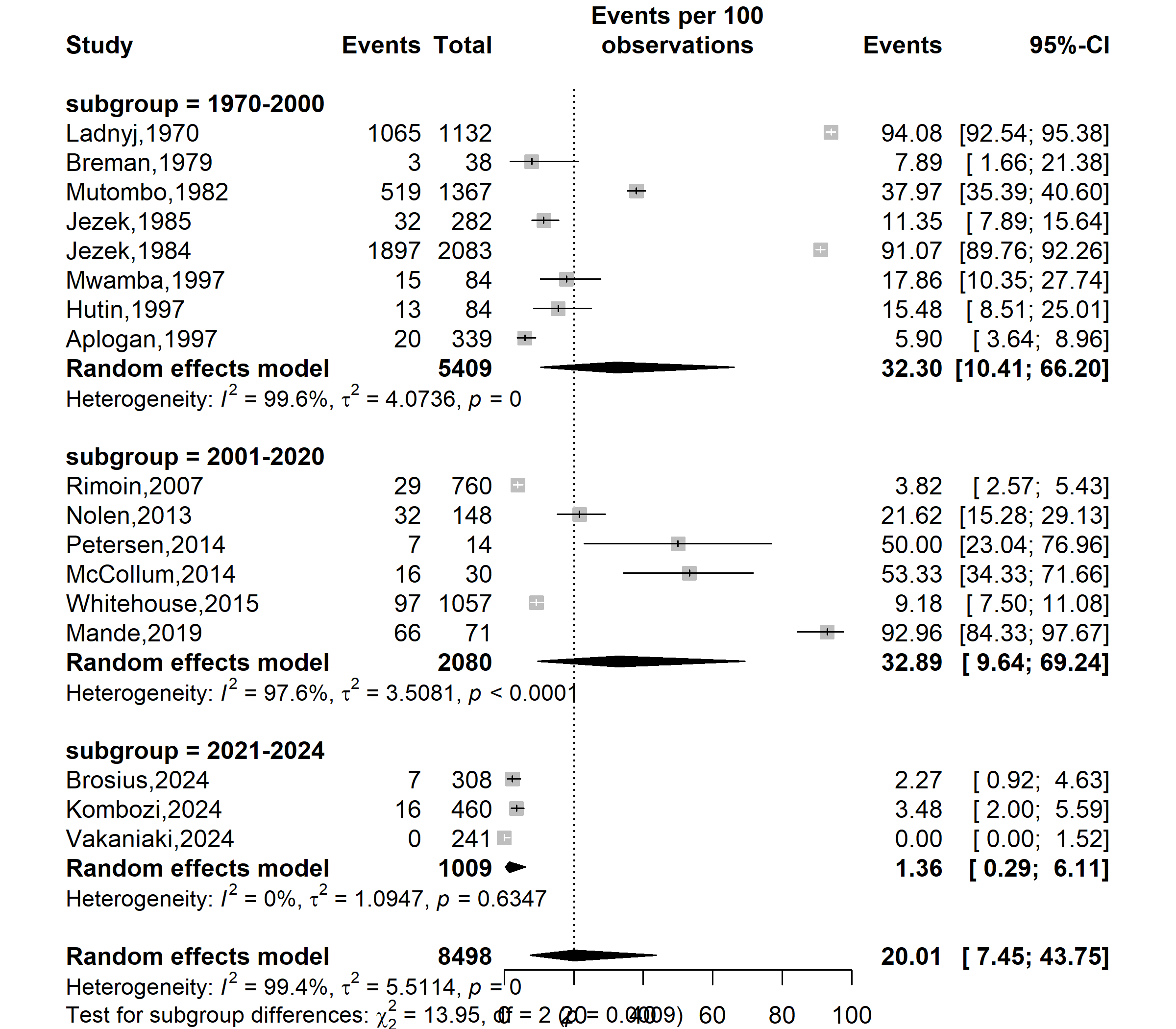

Supplementary Fig. 1 Subgroup analysis assessing the trend of Mpox vaccine uptake pooled estimates in DRC from 1970 to 2024

(1970–2000: smallpox vaccine era; 2001–2020: Limited Mpox vaccine interest; 2021–2024: Increased focus due to global outbreaks)

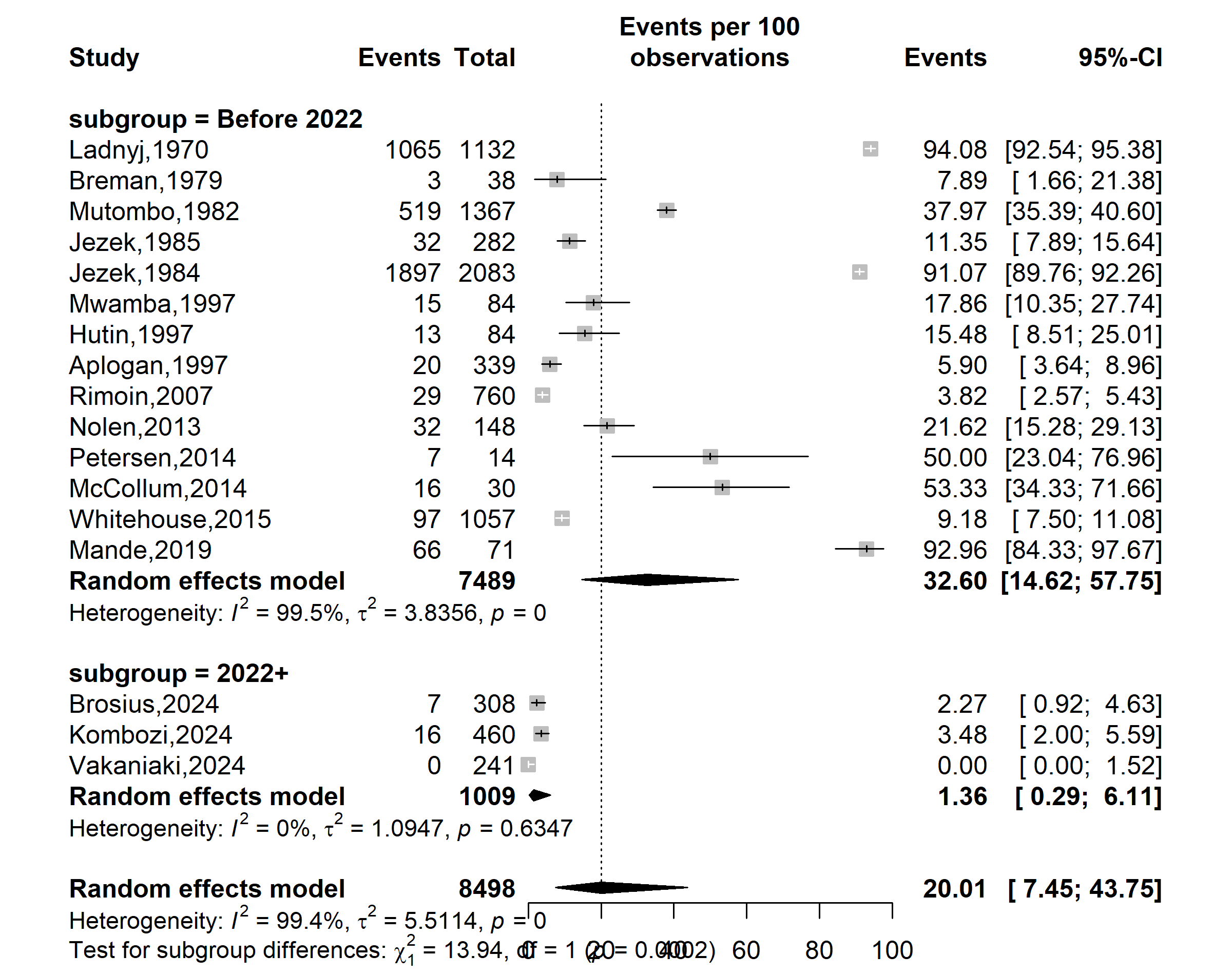

**Vaccine coverage (%)**

**Event rate (%)**

Supplementary Fig. 2 Subgroup analysis assessing of Mpox vaccine uptake pooled estimates before and after the 2022 global pandemic resurgence in DRC

(Period before and after the global outbreak declaration)

Site

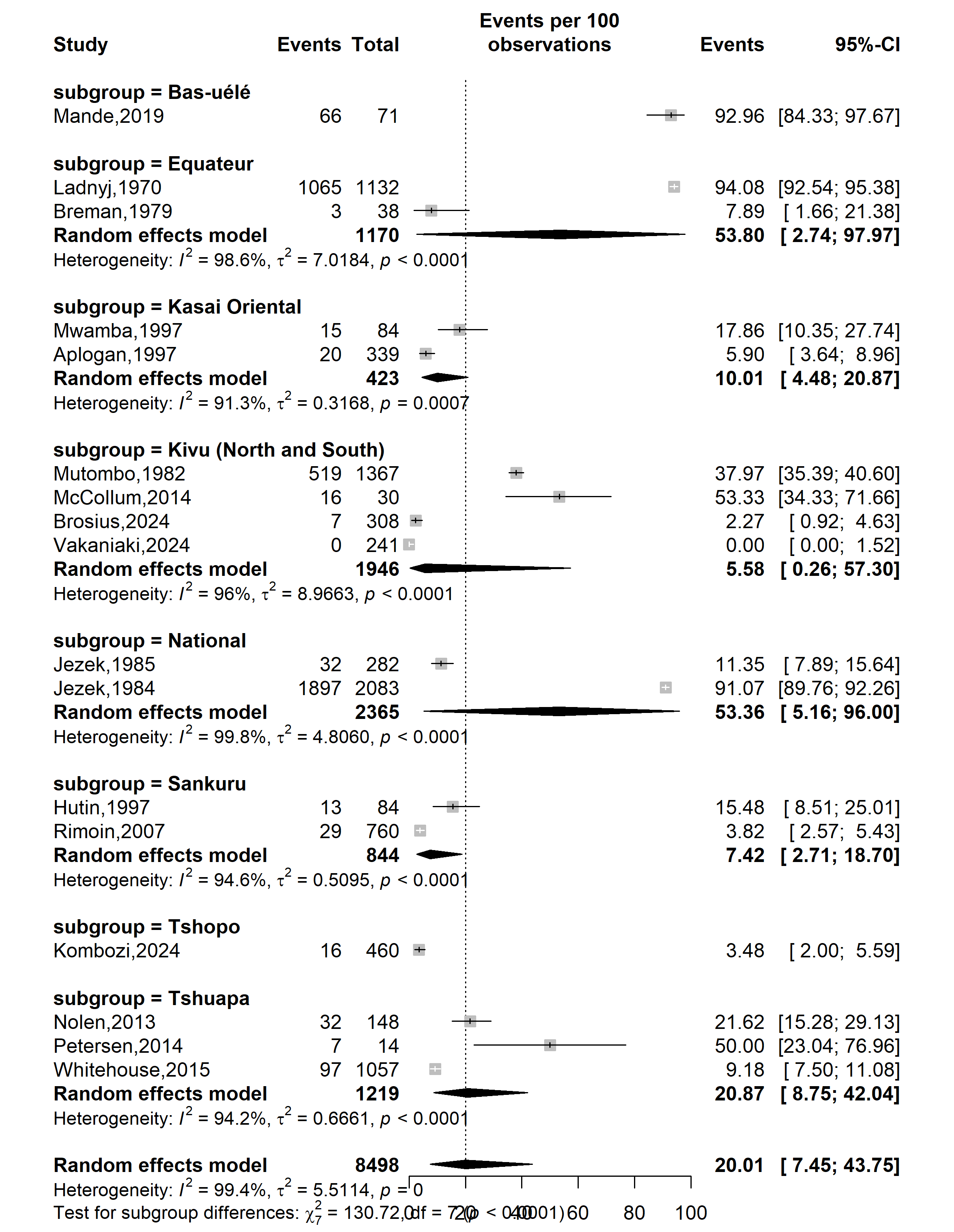

**Vaccine coverage (%)**

**Event rate (%)**

Supplementary Fig. 3 Subgroup analysis assessing geographic disparity of Mpox vaccine uptake pooled estimates in DRC from 1970 to 2024

Study zone

**Vaccine coverage (%)**

**Event rate (%)**

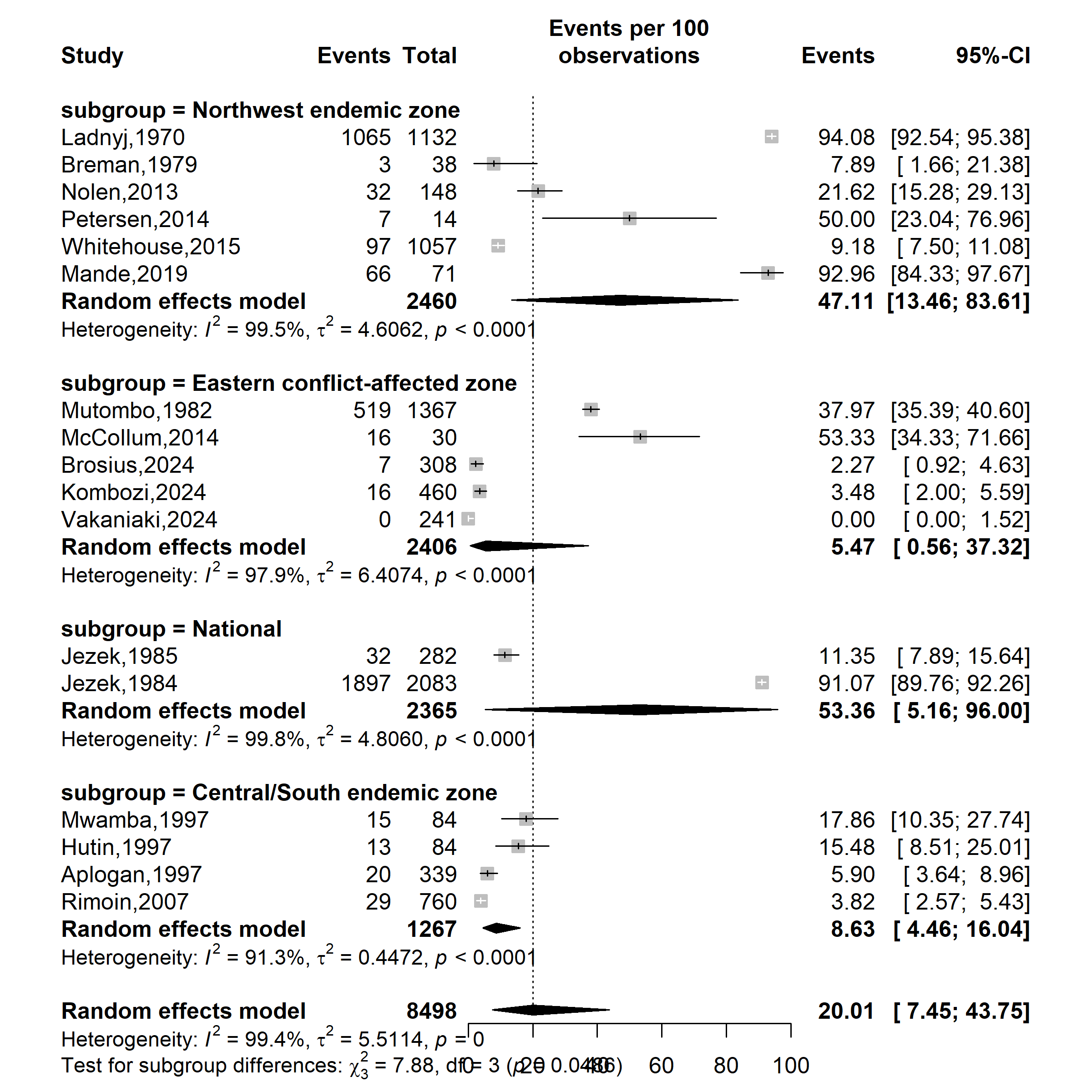

Supplementary Fig. 4 Subgroup analysis assessing disparity of Mpox vaccine uptake pooled estimates in DRC from 1970 to 2024 by geographic zone

(Northwest endemic zone (historically high Mpox transmission zone) = Equateur, Tshuapa and Bas-Uélé; Central/South endemic zone (moderate Mpox activity) = Sankuru and Kasai Oriental; Easthern conflict affected zone (lower Mpox activity but high population movement) = South Kivu, North Kivu and Tshopo)

Setting

**Event rate (%)**

**Vaccine coverage (%)**

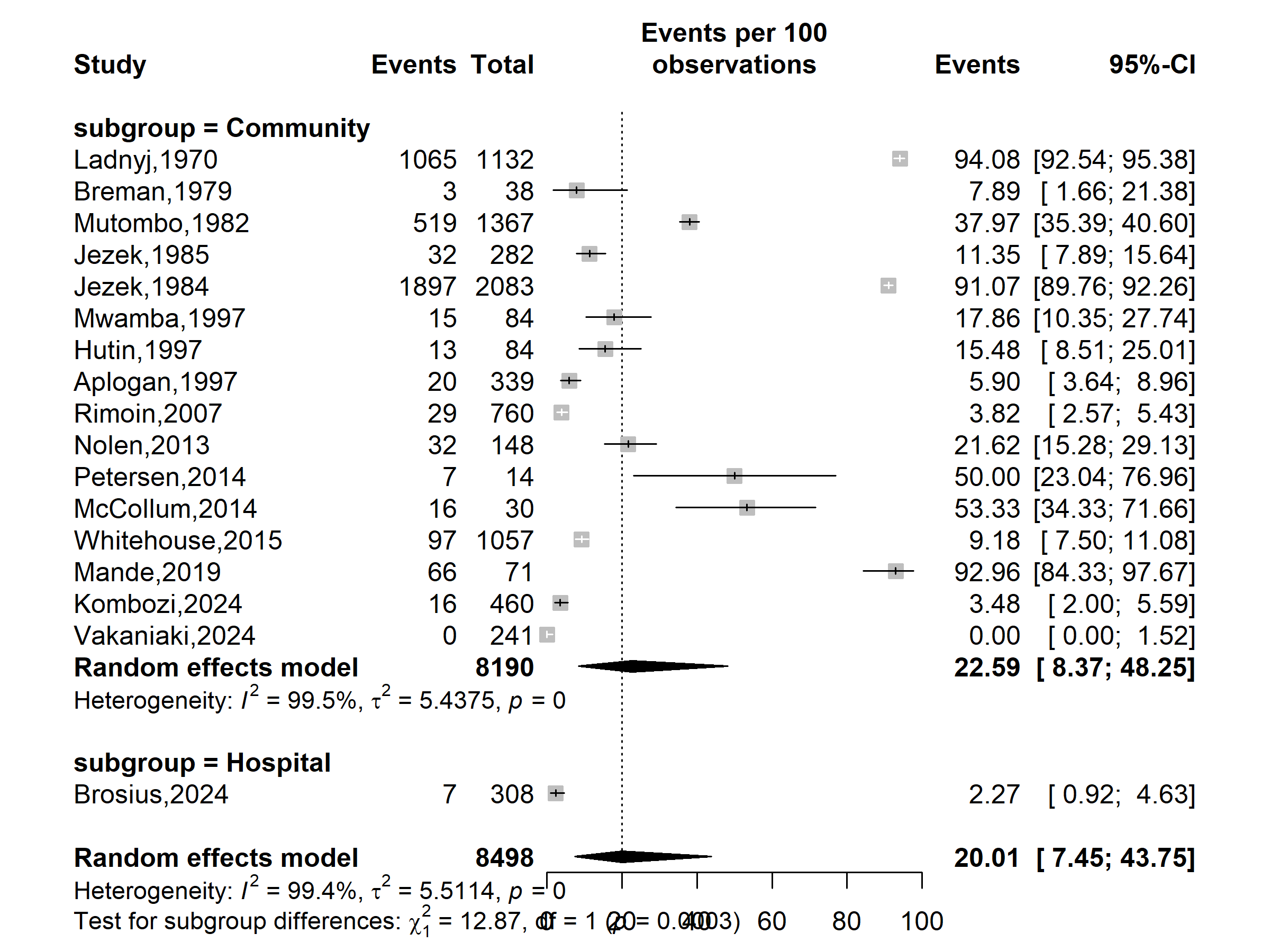

Supplementary Fig. 5 Subgroup analysis assessing the Mpox vaccine uptake pooled estimates according to study setting in DRC from 1970 to 2024

Participant

**Event rate (%)**

**Vaccine coverage (%)**

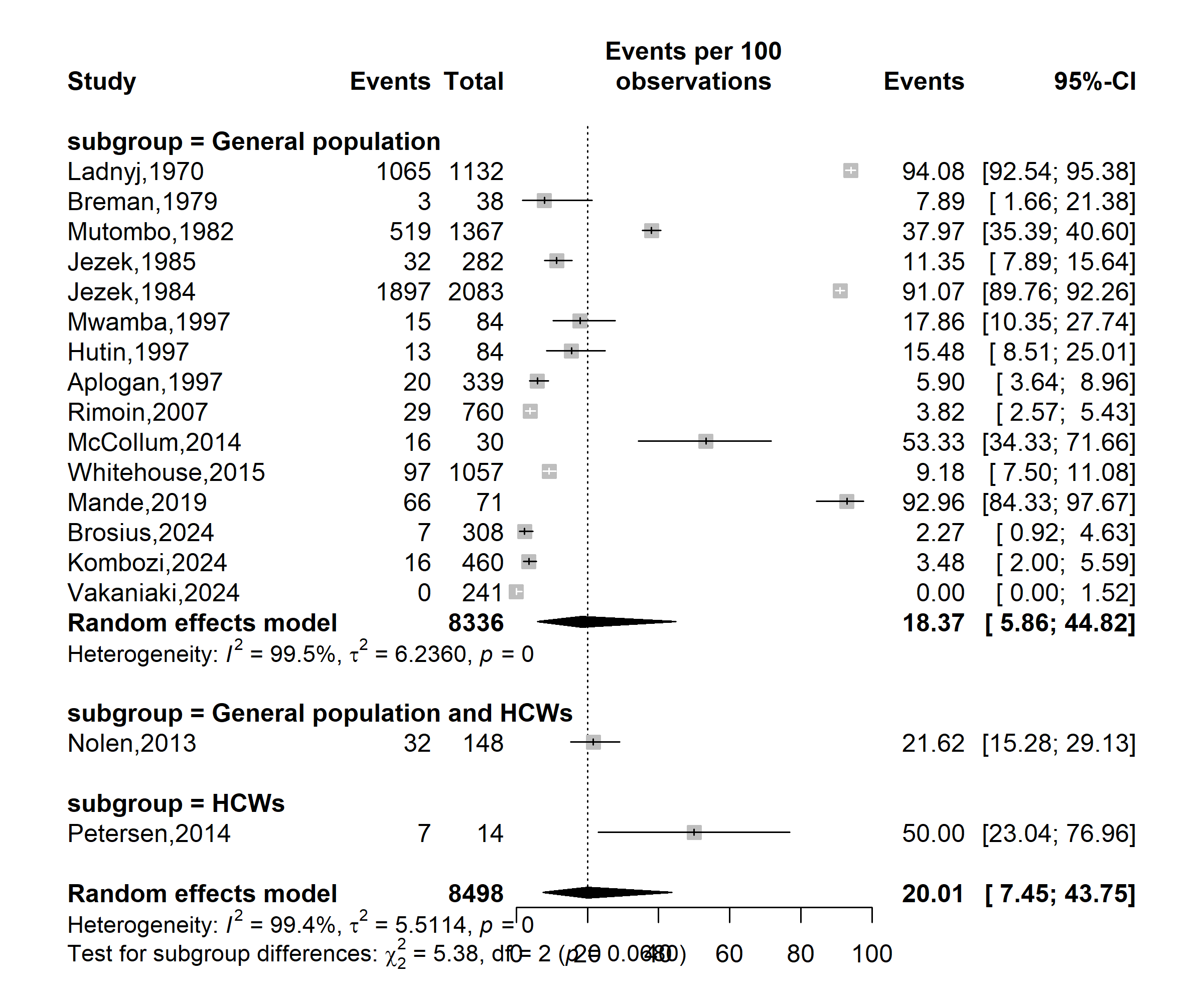

Supplementary Fig. 6 Subgroup analysis assessing the Mpox vaccine uptake pooled estimates according to study participants in DRC from 1970 to 2024
